# Area-based socioeconomic inequalities in cancer mortality in Germany – widening, narrowing or reversing inequalities between 2003 and 2019?

**DOI:** 10.1101/2023.05.16.23290028

**Authors:** Fabian Tetzlaff, Enno Nowossadeck, Lina Jansen, Niels Michalski, Ben Barnes, Klaus Kraywinkel, Jens Hoebel

**Affiliations:** Division of Social Determinants of Health, Department of Epidemiology and Health Monitoring, Robert Koch-Institute, Berlin, Germany; Epidemiological Cancer Registry Baden-Württemberg, German Cancer Research Center (DKFZ), Heidelberg, Germany; German Centre for Cancer Registry Data, Department of Epidemiology and Health Monitoring, Robert Koch-Institute, Berlin, Germany

**Author notes:** **Corresponding author** Fabian Tetzlaff Robert Koch-Institut, FG28 Social Determinants of Health Nordufer 20, 13302 Berlin Germany.

**Keywords:** cancer mortality, time trend, socioeconomic deprivation, GISD, Germany

## Abstract

Cancer mortality has declined in recent decades, but — due to a lack of national individual-level data — it remains unclear whether this applies equally to all socioeconomic groups in Germany. Using an area-based approach, this study investigated socioeconomic inequalities in cancer mortality and their secular trends on a German nationwide scale for the first time. Official cause-of-death data from 2003– 2019 were linked to the district-level German Index of Socioeconomic Deprivation (GISD). Age-standardised mortality rates for all cancers combined and the most common site-specific cancers were calculated according to the level of regional socioeconomic deprivation. To quantify the extent of area-based socioeconomic inequalities in cancer mortality, absolute (SII) and relative (RII) indices of inequality were estimated using multilevel Poisson models. On average, cancer mortality was 50% (women) and 80% (men) higher in Germany’s most deprived than least deprived districts (absolute difference: 84 deaths per 100,000 in women and 185 deaths per 100,000 in men). As declines in cancer mortality were larger in less deprived districts, the socioeconomic gap in cancer mortality widened over time. This trend was observed for various common cancers. Exceptions were cancers of the lung in women and of the pancreas in both sexes, for which mortality rates increased over time, especially in highly deprived districts. Our study provides first evidence on increasing socioeconomic inequalities in cancer mortality on a nationwide scale for Germany. Area-based linkage allows to examine socioeconomic inequalities in cancer mortality across Germany and identify regions with high needs for cancer prevention and control.

## Introduction

Malignant neoplasms are among the leading causes of death, especially in high-income countries, which are most affected by demographic ageing^1, 2^. Numerous studies have shown that a higher socioeconomic position is associated with better health and lower levels of mortality^3, 4^. In terms of cancer, international evidence reveals considerable socioeconomic inequalities in the incidence of various cancers (e.g. ^5-8^)) and survival after cancer diagnosis (e.g. ^9, 10^), as well as in cancer mortality at the population level (e.g. ^7, 11^). These inequalities have been found for many of the most common site-specific cancers^7, 11^. Thus, studies examining socioeconomic inequalities in cancer mortality are highly relevant from a public health perspective.

Studies from various high-income countries report substantial inequalities in cancer mortality and a decline in age-standardised cancer mortality rates over time, with lower mortality levels and steeper mortality reductions in higher socioeconomic groups. This applies to both individual-level (e.g. ^12, 13^) and area-based (e.g. ^7, 11, 14-16^) studies of socioeconomic inequalities in cancer. Comparable evidence for Germany on a nationwide scale is incomplete so far. Due to data restrictions by law, the German death statistics and cancer registries do not contain any information on socioeconomic status, such as education, occupation or income. The few studies available are therefore based either on health insurance data^6, 17^, which contain certain socioeconomic information about the insured individuals, or on regional comparisons using data of the German cause-of-death statistics^18-20^.

First studies for Germany indicate considerable regional and socioeconomic disparities in cancer mortality^6, 17-20^. Recent studies based on German health insurance data report increases in cancer-free life expectancy for total as well as for most of the most common site-specific cancers over time^6, 17^. This development is predominantly driven by decreases in cancer incidence rates, which have been stronger in higher socioeconomic groups. Furthermore, the studies suggest that mortality from cancer differs substantially by socioeconomic position, showing that both incidence risks and survival after cancer diagnosis, as well as the mortality due to causes other than cancer are strongly associated with socioeconomic position^6, 17^. Regional analyses of causes of death, smoking attributable mortality (e.g. lung cancer) and preventable cancer deaths show clear differences between the western and eastern German federal states. These differences still exist 30 years after the German reunification and are associated with socioeconomic inequalities between the two parts of the country. However, the results of the study also suggest that this regional divide may change to a north-south divide with lower cancer mortality in the south than in the north in the future^18-20^. German health insurance data offer the advantage of large case numbers, detailed diagnosis documentation and information on socioeconomic position and mortality of the insurance population at the individual level. However, the data exclude privately insured individuals and do not cover the entire German population. In contrast, regional studies show possible spatial distributions of higher mortality patterns within the entire country, but without the necessary background information about the socioeconomic distribution within Germany, they do not allow conclusions about socioeconomic inequalities in cancer mortality on a nationwide scale. Our study aims to narrow this gap in research and provide first evidence on area-based socioeconomic inequalities in cancer mortality and their trends over time on a nationwide scale for Germany.

Our study is guided by the following research questions:

1. Are there socioeconomic inequalities in cancer mortality in Germany?
2. Do time trends in cancer mortality differ by socioeconomic groups?
3. Have socioeconomic inequalities in cancer mortality decreased (narrowed) or increased (widened) over time?
4. Do socioeconomic differences in cancer mortality and their trends in Germany differ between common cancers?

## Methods

### Data and cause-of-death definition

To analyse socioeconomic inequalities in cancer mortality, we used the annual data of the official German population statistics and cause-of-death (CoD) statistics from 2003 to 2019^21^. The CoD statistics are a full record of all deaths, as the reporting of all deaths in Germany is mandatory. The underlying disease recorded on the death certificate is included in the statistics as the official cause of death, coded according to the ICD-10. The data were aggregated by year, sex, single-year age group and district.

As socioeconomic information of the deceased individuals is not part of this data, we used an area-based approach with spatial data linkage to examine socioeconomic mortality inequalities. At the finest level of geographic aggregation available in the CoD data, which is the 401 German districts, the data are subject to strict legal protections. Therefore, they were accessed on-site at the research data centre of the Federal Statistical Offices. During the period analysed in this study, there have been several district reforms in Germany, in which districts were merged. Therefore, population and CoD statistics were harmonised to the district level of 31.12.2019 to allow for comparisons over time.

We used the same ICD-10 codes to define all cancer and site-specific cancers (table S1, supplemental material) as used in our previous studies^5, 6^ to ensure consistency. Since the analysis focused on primary malignant neoplasms as cause of death, we decided to exclude secondary tumours (C77-79) as well as diagnoses of in-situ neoplasms, benign neoplasms and neoplasms with uncertain or unknown behaviour (D00-D48). Furthermore, we exclude non-melanoma skin cancers (C44) due to incomplete registration of these cancers.

### Regional socioeconomic deprivation

To analyse socioeconomic inequalities in cancer mortality, the cause-of-death statistics were linked with the German Index of Socioeconomic Deprivation (GISD; release 2022v1)^22, 23^ by age, sex, calendar year and district. The GISD is a composite index of area-based indicators in the domains of education, employment and income and is available for the years 1998 to 2019. The index combines the three dimensions with equal weight. The scores for the dimensions are calculated based on separate principal component analyses with three indicators for each dimension (employment: unemployment and employment rate, gross wage and salary; income: net household income, debtor quota, tax revenue; education: the proportion of: employees with university degree, employees without qualification, school leaving without qualification)^22^. The indicators are obtained from the INKAR database (Indicators, Maps and Graphics on Spatial and Urban Monitoring) of the Federal Institute for Research on Building, Urban Affairs, and Spatial Development^24^ and statistics of the Federal Employment Agency^25^. For our analysis of time trends, the districts were assigned to quintiles according to their GISD score and were later categorised into three groups (low=Q1; middle=Q2 to Q4; high=Q5).

Further information and a more detailed methodical description of the GISD score and its calculation can be found in the recently published revision of the German Index of Socioeconomic Deprivation in Michalski et al.^22^. The GISD is published under CC by 4.0 licence and is freely accessible via the GitHub website of the Robert Koch-Institute (https://github.com/robert-koch-institut or https://doi.org/10.5281/zenodo.6840304).

### Statistical analysis

First, to allow for a general overview of inequalities and time trends in cancer mortality we calculated the mortality rate for all cancers combined over the entire study period and the annual mortality rates for the most common cancers, stratified by sex and levels of socioeconomic deprivation. Mortality rates were calculated as age-standardised mortality rates (deaths per 100,000 population; European Standard Population 2013^26^).

Second, to quantify the extent of absolute und relative area-based socioeconomic inequalities in cancer mortality, the regression-based Slope Index of Inequality (SII) and Relative Index of Inequality (RII) were estimated using multilevel Poisson regression models. These measures are frequently used to investigate the magnitude of absolute and relative health inequality^27, 28^. In contrast to simple two-groups comparisons of rate differences and rate ratios (e.g. by deprivation quintiles), SII and RII consider the entire socioeconomic spectrum in determining socioeconomic inequalities in the outcome variable^27^. Accordingly, the average mortality risk is compared between the districts at the lowest and highest levels of the regional socioeconomic distribution, considering the risk of the districts in between by using a regression-based estimation approach. The regression models used to estimate these summary measures of health inequality were stratified by sex and calendar year. The models contained age as age-groups (0-59 years and 5-year age groups up to 85+ years) and the district-level GISD-score ranging from 0 (least deprived) to 1 (most deprived) as predictor variables. The models were fitted as multilevel models with age groups as first-level and districts as second-level units. To allow for comparisons over time, the models were weighted with the 2013 European Standard Population. We used the logarithm of the population size in each age group as offset term in the regression models to account for the varying population at risk in each district. The analyses were performed with STATA 17 and R 4.1.2.

## Results

Over the entire observation period of 17 years, an annual average of nearly 82 million persons at risk of dying from cancer were included in the analyses. The least deprived quintile of districts had the highest, and the most deprived quintile the lowest number of person-years. Across all cancer types, an average of approximately 219,000 deaths per year (101,000 women and 118,000 men) were recorded between 2003 and 2019 (table 1).

**Table 1.**
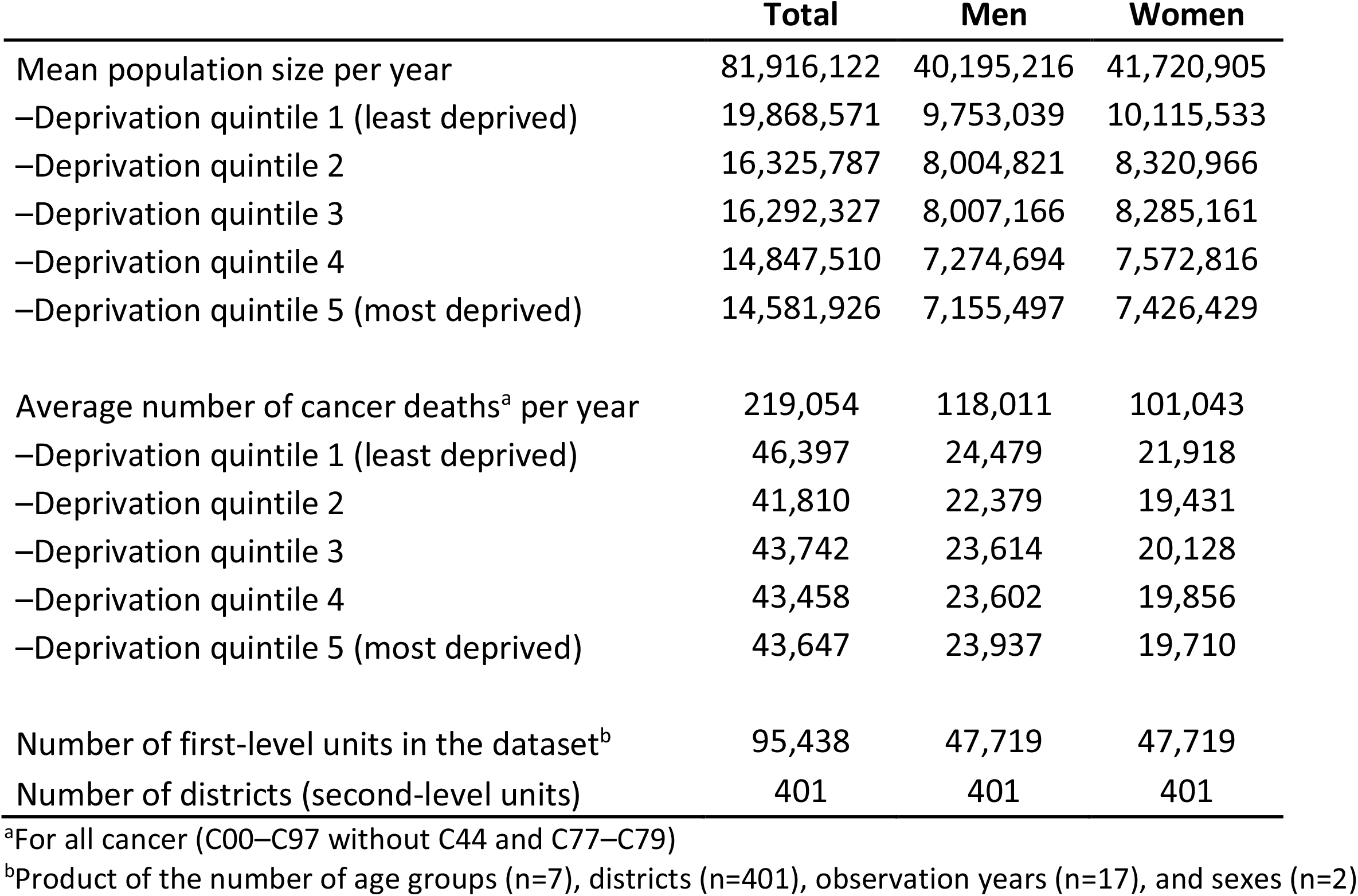
Description of the study population, 2003–2019

### Area-based socioeconomic inequalities in cancer mortality

Figure 1 displays the age-standardised mortality rate for all cancers combined between 2003 and 2019 and its area-based socioeconomic distribution across deprivation quintiles. For both sexes, there was a considerable socioeconomic gradient in cancer mortality over the entire period, with increasing mortality rates with higher levels of regional socioeconomic deprivation. The average annual age-standardised cancer mortality rate among women living in the least deprived quintile of districts was 198 deaths per 100,000 population, while the average annual rate for women of the most deprived quintile was 215 deaths per 100,000 population (Q1=198; Q2=202; Q3=208; Q4=210; Q5=215). Among men, the average annual age-standardised cancer mortality rate overall was substantially higher than among women, and the area-based socioeconomic gradient was more pronounced (Q1=301; Q2=318; Q3=336; Q4=346; Q5=372). Across the entire range of the GISD score, this led to an absolute difference between the least and most deprived districts of an average of 84 cancer deaths per 100,000 women and 185 cancer deaths per 100,000 men per year. From a relative perspective, a 50 percent higher cancer mortality among women (RII=1.5) and an 80 percent higher cancer mortality among men (RII=1.8) was observed comparing most to least deprived districts in Germany.

**Figure 1.**
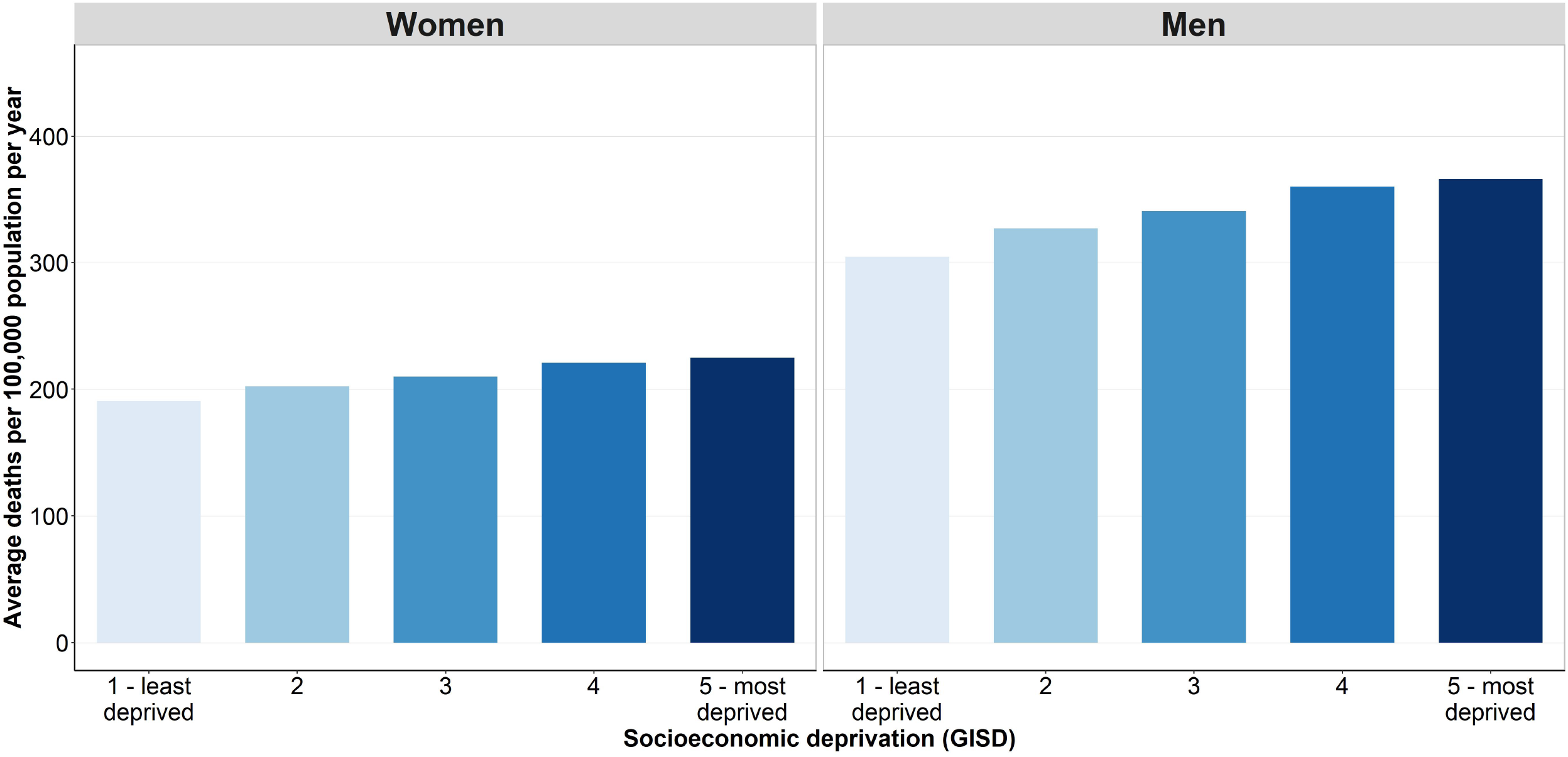
Age-standardised cancer mortality by quintiles of regional socioeconomic deprivation and sex, 2003–2019

### Time trends in cancer mortality

Figure 2 displays the development of age-standardised cancer mortality by regional socioeconomic deprivation over time. For men, cancer mortality decreased in all socioeconomic groups, with a larger decline in less deprived districts. Among women, cancer mortality decreased in districts with middle and low socioeconomic deprivation, whereas it remained largely unchanged in districts with high deprivation. The socioeconomic gap in mortality from all cancers combined among women was not apparent at the beginning of the period and only became apparent in later years.

**Figure 2.**
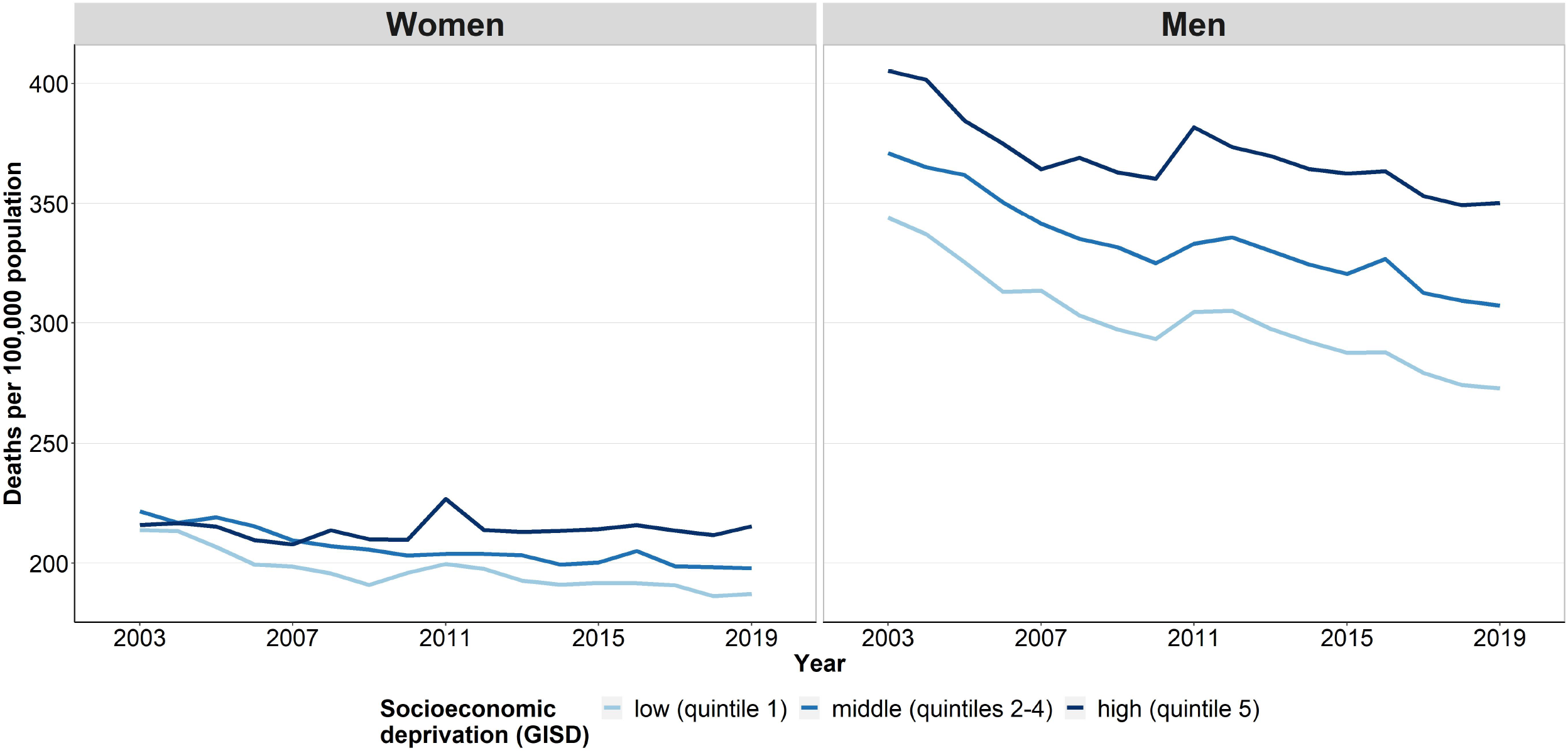
Time trends in age-standardised cancer mortality by sex and regional socioeconomic deprivation, 2003–2019

For almost all site-specific cancers considered, the standardised mortality rates declined but inequalities persisted or increased with higher levels of cancer mortality in more deprived districts (figure 3 and figure S1, supplemental material). This applies to cancers of the colon and rectum, stomach, kidney and bladder in both sexes and lung cancer in men, as well as to the sex-specific cancers of the prostate, ovary and cervix, for which mortality rates also declined in all groups of regional socioeconomic deprivation (figure 3 and figure S1, supplemental material). However, female breast cancer mortality decreased only in districts of low and middle socioeconomic deprivation. Although breast cancer mortality in districts with high deprivation was initially at a lower level than in districts with middle and low deprivation, it stagnated or even tended to increase over time (figure 3). For cancers of the oesophagus, the lymphatic and haematopoietic system in both sexes and liver cancer in women, age-standardised mortality rates also stagnated irrespective of level of regional socioeconomic deprivation, with consistently higher mortality in more deprived districts. In contrast, male liver cancer mortality increased, especially in districts with high and middle socioeconomic deprivation. Increases in mortality rates were also found for lung cancer among women and cancers of the pancreas, oral and upper respiratory tract in both sexes irrespective of the level of socioeconomic deprivation, with higher mortality rates in more deprived districts in almost all years of observation. The fluctuations caused by the low number of annual deaths from malignant melanoma did not allow for a clear interpretation of the trend and patterns of socioeconomic inequalities in standardised mortality rates (figure S1, supplemental material).

**Figure 3.**
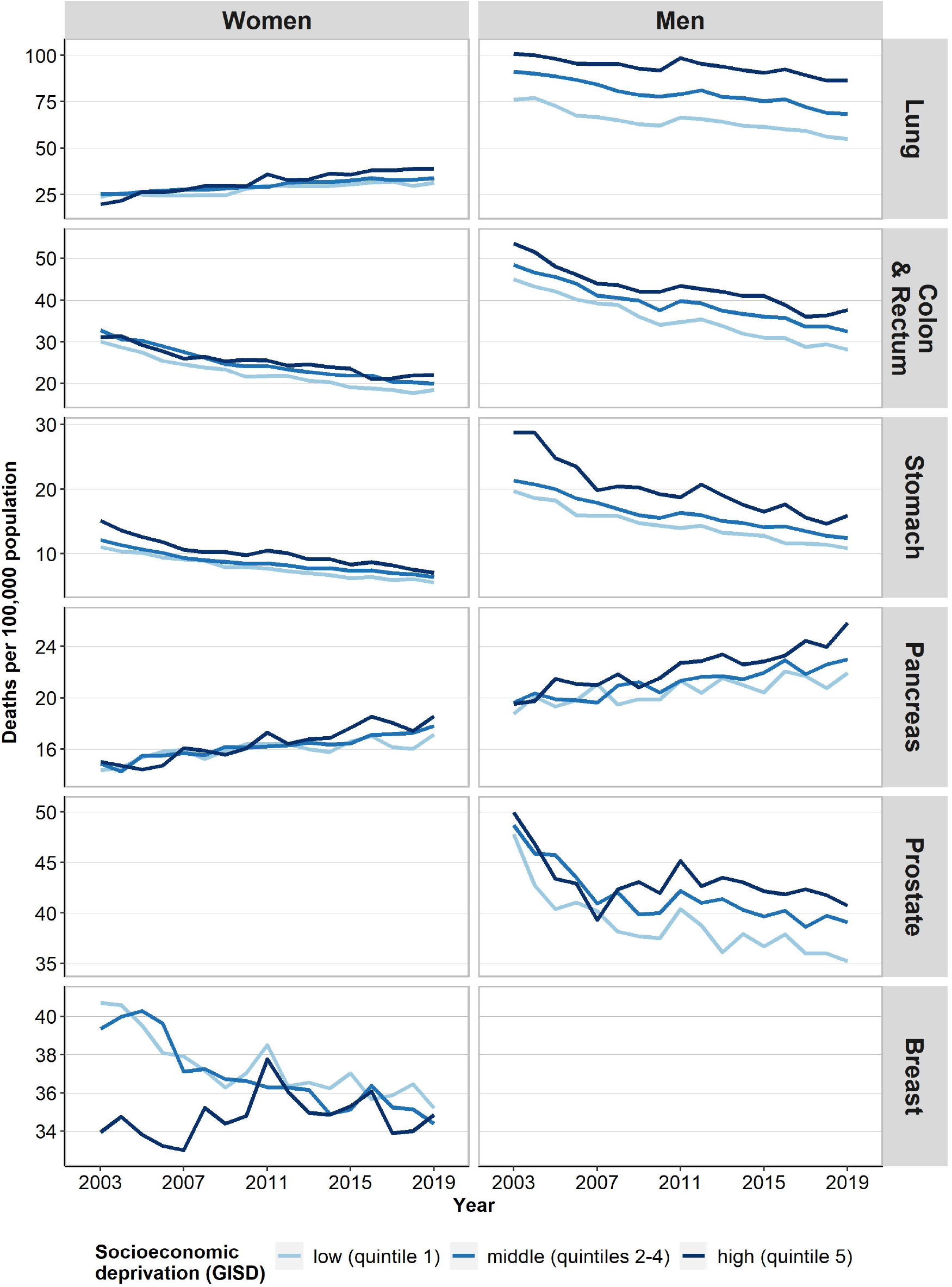
Time trends in age-standardised mortality from the most common cancers by sex and regional socioeconomic deprivation, 2003–2019

### Time trends in absolute and relative inequalities

Considering the SII and RII for all cancers combined over time, the mortality gap between the most und least deprived districts widened substantially in both sexes (figure 4). This was also true for the majority of the most common site-specific cancers (figure 5). We found clear increases in absolute and relative inequalities in mortality from lung, colon and rectum, and pancreas cancer in both sexes as well as in prostate cancer. Inequalities increased particularly in lung cancer, where, for instance, the RII for women almost quadrupled over time. The development was even stronger for women than for men. In female breast cancer mortality, we found reverse absolute and relative inequalities before the year of 2010, with higher mortality rates in the least than most deprived districts. From 2010 onwards, the socioeconomic gradient in female breast cancer mortality turned to higher mortality rates in the most compared to the least deprived districts (figure 5). In all other specific cancers considered, patterns were similar to the development of SII and RII in all cancers combined (figure S2, supplemental material). However, cancers of the oral and upper respiratory tract, oesophagus, liver, kidney, and lymphatic and haematopoietic system showed stronger increases in absolute and relative mortality inequalities in men than in women. This resulted in widening differences in area-based socioeconomic inequality in cancer mortality between men and women in both absolute and relative terms (figure S2, supplemental material). Moreover, we found a reversing socioeconomic gradient for malignant melanoma in both sexes, indicating that after 2009 (women) and 2010 (men), malignant melanoma mortality was higher in the most deprived districts than in the least deprived districts (figure S2, supplemental material).

**Figure 4.**
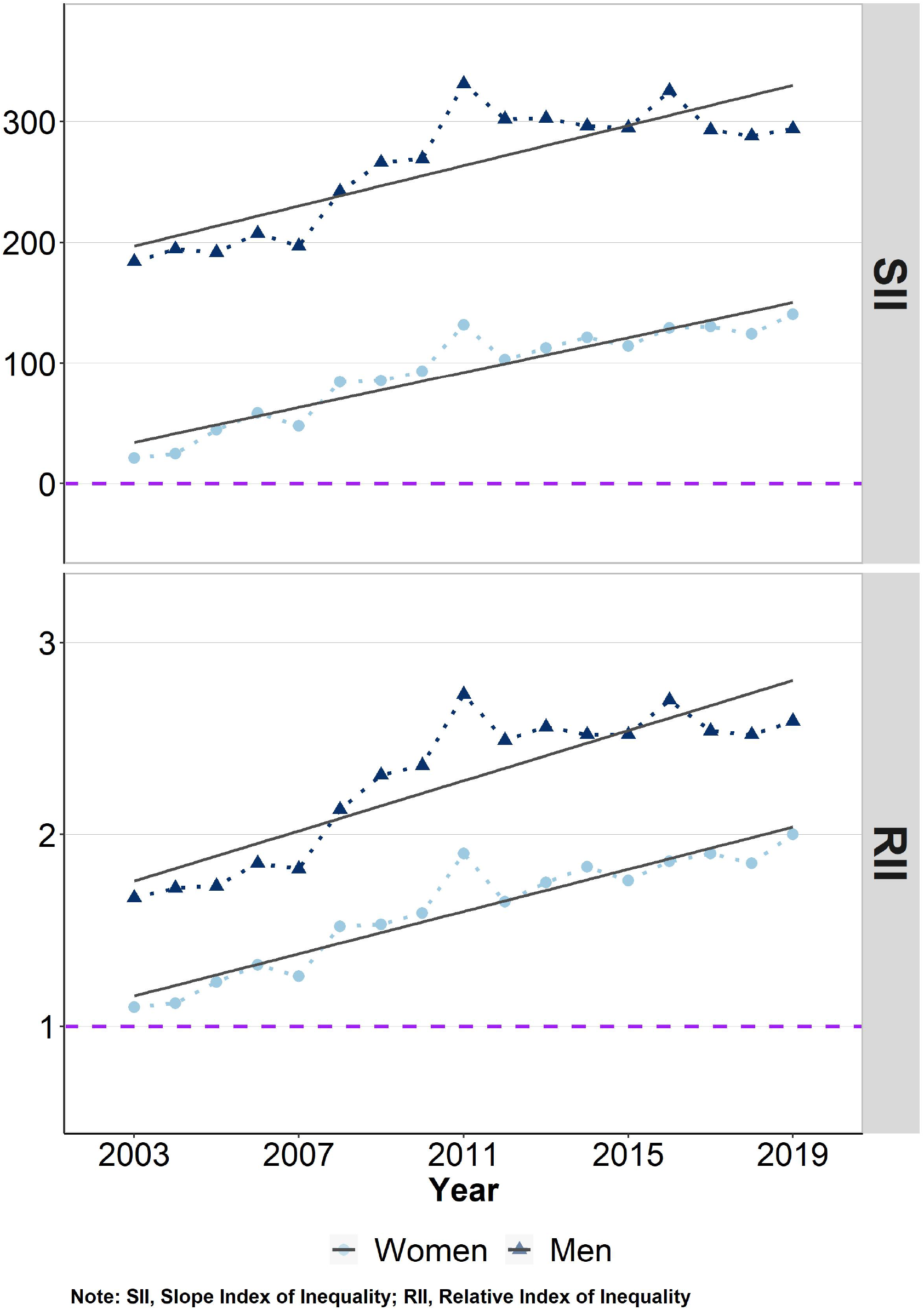
Absolute (SII) and relative (RII) inequalities in age-standardised cancer mortality by sex and year, 2003–2019 Note: SII, Slope Index of Inequality; RII, Relative Index of Inequality

**Figure 5.**
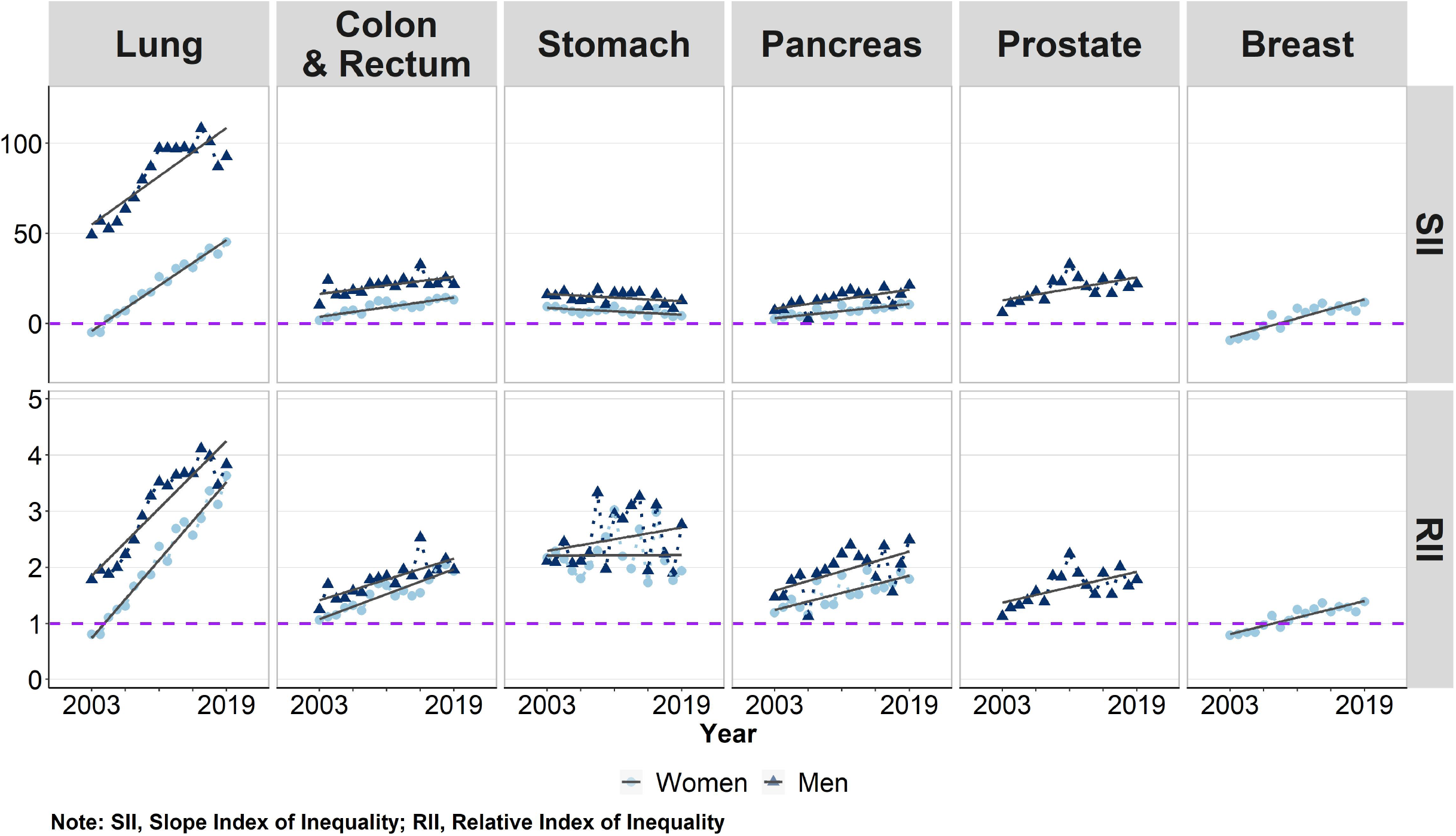
Absolute (SII) and relative (RII) inequalities in age-standardised mortality from the most common cancers by sex and year, 2003–2019 Note: SII, Slope Index of Inequality; RII, Relative Index of Inequality

## Discussion

This is the first study linking small-area data from the official German cause-of-death statistics with a regional socioeconomic deprivation index to analyse socioeconomic inequalities in cancer mortality on a German nationwide scale. Our analyses have shown that there are pronounced area-based socioeconomic inequalities in cancer mortality overall as well as in mortality from the most common site-specific cancers to the disadvantage of people living in Germany’s socioeconomically more deprived districts. Moreover, the study shows that these inequalities widened over time for all cancers combined as well as for most of the analysed site-specific cancers.

The positive trend of overall declining cancer mortality is primarily due to the clear decrease in mortality from colorectum, and stomach cancer in both sexes, as well as from lung and prostate cancer in men and breast cancer in women. Since the decline in cancer mortality was strongest in districts with low and middle socioeconomic deprivation, these regions contributed most to this development, while highly deprived regions experienced the smallest declines in cancer mortality. The increase in pancreatic cancer mortality in both sexes and lung cancer mortality in women in all socioeconomic groups, although concerning, had only a small impact on total cancer mortality. These negative trends in site-specific cancer mortality and the trend towards increasing area-based socioeconomic inequalities in cancer mortality should be investigated in additional studies. This can be an important step towards a better understanding of the relationship between socioeconomic deprivation and mortality, and to identify opportunities to reduce cancer mortality and improve health equity.

Similar to other high-income countries^7, 11, 14-16^, our study found higher mortality rates in the socioeconomically most deprived districts. This is true for almost all cancers considered. In Germany, the majority of the most deprived districts are located in the eastern German Federal States^22^. The only exceptions here are the larger east German cities (e.g. Jena, Dresden, Berlin), which are less deprived compared to the German average deprivation level^22^, and consequently have a lower cancer mortality. Studies for Germany examining inequalities in cancer survival indicate that the composition of cancers can differ considerably between deprivation groups with respect to the distribution of site-specific cancers. Thus, cancers that have a much better overall survival prognosis and cancers with an earlier stage at diagnosis are more often represented in the lower deprivation group^9, 29^, which have an impact on overall cancer mortality and on increasing inequalities in all cancers combined over time.

As in most European countries^11, 14^, our study found the strongest socioeconomic inequalities in lung cancer. This was true for both sexes (at least at the end of the study period), although the inequalities were much larger among men. A negative association between cancer mortality and regional socioeconomic deprivation was found for breast cancer and lung cancer in women at the beginning of the observation period in the early 2000s. For both cancers, the socioeconomic gradient reversed over time, so that in the recent past, the highest mortality rates were found in the districts with the highest level of socioeconomic deprivation as with all other cancers considered. This is in line with a current study which also reported a gradient reversal for lung cancer mortality among women in Estonia, Slovenia and Italy^11^. In Germany, similar results were reported for lung cancer incidence and cancer-free life expectancy^6, 17, 30^. The close association between the incidence and mortality of lung cancer suggests that as incidence decreases, mortality in men should continue to decrease, while in women mortality should increase due to rising incidence. Our results support this assumption as mortality from lung cancer decreased in men but increased in women.

We found the largest socioeconomic inequalities in mortality from cancers (e.g. lung cancer, colon and rectum, stomach cancer, etc.), which are strongly associated with avoidable risk factors (e.g. smoking, alcohol consumption, severe obesity, etc.)^18^. Tobacco consumption is the main risk factor for lung cancer and the development of lung cancer mortality reflects the state of the smoking epidemic^31^. Germany can today be assigned to the fourth phase of the smoking epidemic with a decreasing lung cancer mortality rate among men and a still increasing lung cancer mortality rate among women. Pampel et al. propose to combine the smoking epidemic concept with the diffusion hypothesis to allow a more precise description of the influence of socioeconomic status on the development of the smoking epidemic^31^. Rogers postulated in the diffusion hypothesis that people with higher socioeconomic status are the first in the population to adapt social innovations^32, 33^. This is true for tobacco, which was considered a luxury good at the beginning of the 20th century, as well as for preventive health knowledge and measures^31-33^. According to Pampel et al., people with a higher socioeconomic position smoked more frequently and died more often from lung cancer with a time lag of one to two decades at the beginning of the smoking epidemic. However, smoking prevention, abstinence and cessation were also first adopted by individuals with a higher socioeconomic position. Consequently, smoking-related mortality decreased most strongly in these groups, followed by more deprived groups, but with a considerable time lag^31^. Our results support this hypothesis: Over the entire period, lung cancer mortality decreased in men in all socioeconomic groups. Among women, in contrast, the mortality rate increased, especially among the socioeconomically highly deprived.

Probably caused by the different tempo of the adaptation of preventive behaviours, such as never-smoking and smoking cessation^34^, the inequalities widened over time. In addition, a reversal of the socioeconomic gradient was observed in lung cancer mortality among women at the beginning of the 2000s, as found in our study. Compared to other European countries, Germany is ranked at the bottom in terms of the comprehensiveness of tobacco-control policies^35^, which leaves much potential to improve smoking prevention and to reduce smoking-related cancer mortality.

However, in addition to tobacco use, there are also other risk factors which may substantially increase the risk of cancer incidence and mortality, such as alcohol consumption (e.g. ^36, 37^), high levels of Ultra Violet radiation exposure (e.g. ^37, 38^), poor housing conditions and high levels of air pollution in the workplace (e.g. ^37, 39^). Improved structural prevention and education, especially in lower socioeconomic groups and their living and working environment, could lead to a reduction in risk factors and thus considerably reduce the risk of cancer and mortality from various cancer entities.

In terms of cancer mortality, it is crucial that tumours are detected at the earliest possible stage^40, 41^. However, there are also clear social inequalities in the stage at diagnosis, with higher and more advanced stages among the most deprived individuals^41, 42^. The preventive cancer screening examinations have a key function in this context. They can contribute not only to reduce the staging at diagnosis and thus the severity of the disease, but also to improve the chances of cancer survival and contribute to reduce inequalities in cancer morbidity and mortality. In Germany, national screening programmes for breast cancer, malignant melanoma, prostate cancer and colorectal cancer have been introduced nationwide since the beginning of the 2000s. The costs for the screening are completely covered by the statutory health insurance funds^43^. However, the screening programmes are not used equally by all socioeconomic groups^43-46^. The socioeconomic unequal utilisation of cancer screening may partly explain inequalities in cancer mortality. Our analyses show that area-based socioeconomic inequalities in mortality from colorectal cancer and malignant melanoma, as well as sex-specific cancers (breast cancer, prostate cancer, ovarian cancer, cervix uteri) covered by national screening programmes have widened considerably. Moreover, the strong reductions in breast cancer mortality found in our analysis may be an indication for the potential success of the nationwide introduction of breast cancer screening, but further improvements seem achievable, especially in deprived populations. Further analyses on inequalities in the utilisation of preventive measures and staging at diagnosis could gain more insights into these aspects and underlying mechanisms.

### Strengths and Limitations

Since the notification of all deaths in Germany is mandatory, complete cause-of-death statistics were available and included in our analysis. Deaths have been recorded continuously, completely, with residential information and according to the ICD-10 coding, which allowed us to analyse the spatio-temporal development of cancer mortality for the entire German population (> 81 million persons at risk). The availability of the cause-of-death statistics at district level made it possible to link the data with an area-based deprivation index to examine socioeconomic inequalities in cancer mortality in Germany at full-population level for the very first time.

The main limitation of the data is that they do not include information on socioeconomic position at the individual level. Therefore, the area-based deprivation index was used to approximate socioeconomic position via the district level. In this respect, it should be borne in mind that this may lead to ecological fallacies. Although merging official mortality data with area-based socioeconomic indices allows the analysis of inequalities in mortality, this type of analysis is likely to underestimate inequalities compared to individual-level analyses, i.a. as the population within a district is heterogeneous in terms of its socioeconomic characteristics. Another limitation of the data is that only the documented underlying disease from the death certificates is included in the cause-of-death statistics. However, due to coding practices that deviate from the defined standard, acute events may be coded more frequently than the actual underlying disease that ultimately led to death. As a result, especially chronic diseases may be under-recorded as causes of death. Considering the entire causal chain in deaths statistics with several causes of death (e.g. multi-causal cause-of-death statistics) could help to prevent this limitation of the current cause-of-death statistics. The possibility that cancer is underestimated as a cause of death cannot be completely ruled out. However, since cancers are usually serious diseases that require extensive therapy, treatment and constant monitoring, a potential underestimation due to coding a cause of death other than cancer on the death certificate may be negligible.

## Conclusion

As the first study investigating socioeconomic inequalities in cancer mortality on a nationwide scale for Germany, our study demonstrates higher risks of cancer death among residents in the country’s more deprived districts. For almost all cancers considered, the mortality rate has declined substantially since the early 2000s but, on average, residents in highly deprived districts experienced the smallest declines. Exceptions are cancers of the lung in women and of the pancreas in both sexes, for which mortality rates increased over time, especially in highly deprived districts. As a result, the socioeconomic gap in total cancer mortality widened in both absolute and relative terms. Since the particular drivers of this development are still not well understood, further investigations should examine the underlying mechanisms in more detail. Overall, the findings suggest that more and effective prevention efforts are needed to reduce cancer mortality in socioeconomically disadvantaged groups. Evidence on area-based socioeconomic inequalities in cancer mortality can help to identify groups and districts with high potential for cancer prevention and control. Tailored public health interventions should be strengthened to reduce cancer mortality in socioeconomically disadvantaged districts in Germany.

### Data Availability Statement

The official German cause-of-death data analysed in this study were provided by the Research Data Centre of the German Statistical Offices. At the lowest available level of area aggregation, the data are subject to strict data protection by law and are not publicly available. The cause-of-death data are available upon request and application to the Research Data Centre of the Statistical Offices.

The German Index of Socioeconomic deprivation is published under CC by 4.0 licence and is freely accessible via the GitHub website of the Robert Koch Institute (https://github.com/robert-koch-institut).

## Data Availability

The official German cause-of-death data analysed in this study were provided by the Research Data Centre of the German Statistical Offices. At the lowest available level of area aggregation, the data are subject to strict data protection by law and are not publicly available. The cause-of-death data are available upon request and application to the Research Data Centre of the Statistical Offices.
The German Index of Socioeconomic deprivation is published under CC by 4.0 licence and is freely accessible via the GitHub website of the Robert Koch Institute (https://github.com/robert-koch-institut).

https://github.com/robert-koch-institut

## Abbreviations

GISD: (German Index of Socioeconomic Deprivation)
SII: (absolute index of inequality)
RII: (relative index of inequality)
CoD: (cause-of-death)
INKAR: (Indicators, Maps and Graphics on Spatial and Urban Monitoring)

## Acknowledgments

We would like to thank the colleagues from the Berlin-Brandenburg Statistical Office for their advice and support.

## Funding

The study was funded by the German Cancer Aid [Deutsche Krebshilfe]. The funder was not involved in the study design, collection, analysis, interpretation of data, the writing of this article or the decision to submit it for publication.

## Author Contributions

FT: Conceptualisation, Data Curation, Formal Analysis, Investigation, Methodology, Project Administration, Visualization, Writing – original draft; EN: Conceptualisation, Writing – review & editing; LJ: Conceptualisation, Writing – review & editing; NM: Conceptualisation, Writing – review & editing; BB: Writing – review & editing; KK: Writing – review & editing; JH: Fundraising, Conceptualisation, Formal Analysis, Methodology, Supervision, Writing – review & editing

## Disclosure of Conflict of Interest

The authors declared no potential conflicts of interest.

## Notes

### Competing Interest Statement

The authors have declared no competing interest.

