## Supplemental Material for "Area-based socioeconomic inequalities in cancer mortality in Germany – widening, narrowing or reversing inequalities between 2003 and 2019?"

***Corresponding author**

**Table S1** Definition of total cancer and site-specific cancers according to ICD-10GM classification

| | |
|---|---|
| total cancer | C00-C97, without C44, C77-79 |
| oral and upper respiratory tract | C00-06, C09-14, C32 |
| oesophagus | C15 |
| stomach | C16 |
| colon | C18-20 |
| liver | C22 |
| pancreas | C25 |
| lung | C33-34 |
| malignant melanoma of skin | C43 |
| (female) breast | C50 |
| cervix uteri | C53 |
| ovary | C56 |
| prostate | C61 |
| kidney | C64 |
| bladder | C67 |
| lymphoid and hematopoietic neoplasms | C81–96 |

### Figure S1 Time trend in age-standardised mortality rate by sex and regional socioeconomic deprivation

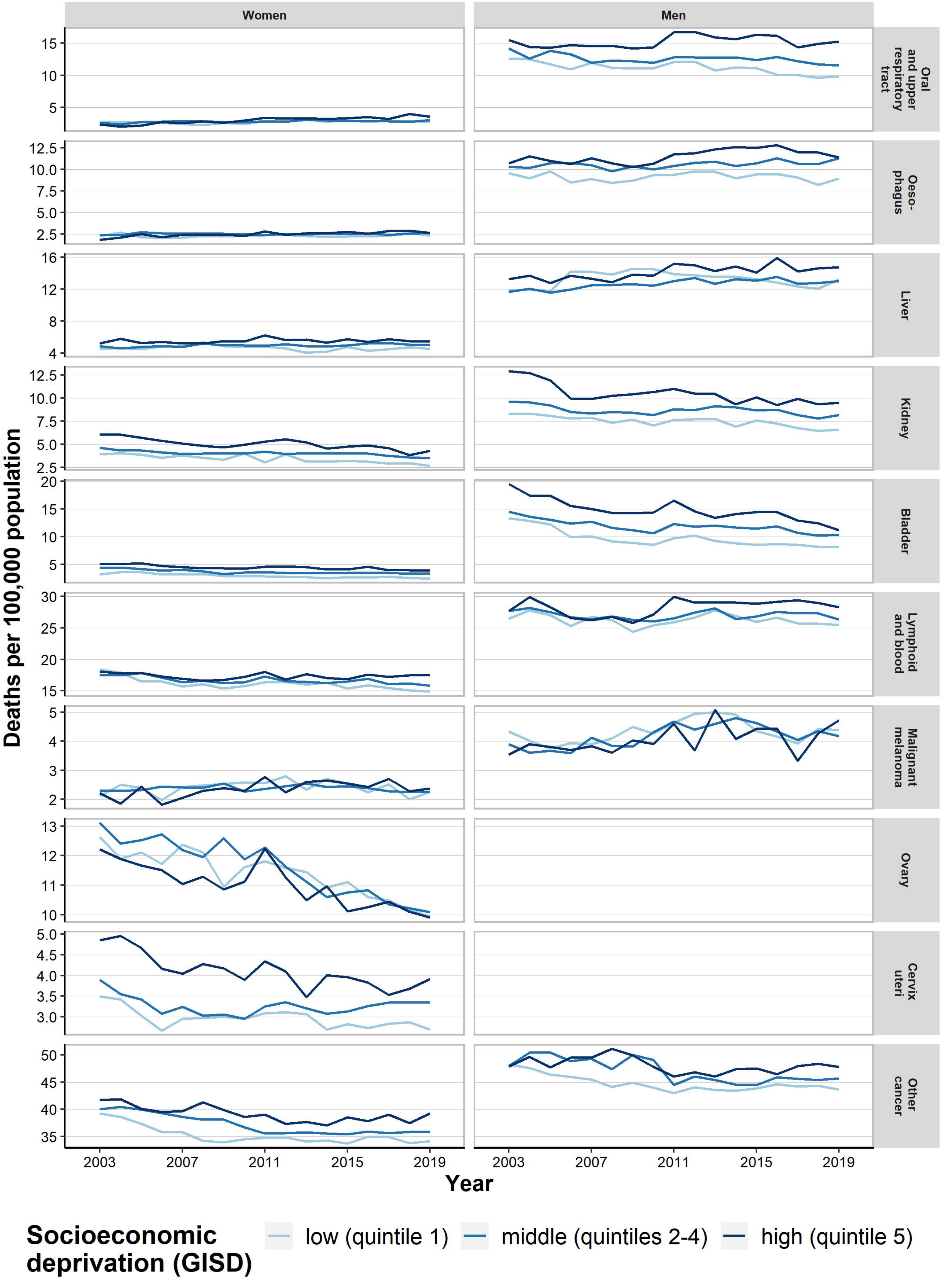

**Figure S2 Absolute (SII) and relative (RII) inequalities in cancer site-specific Mortality by sex and year**

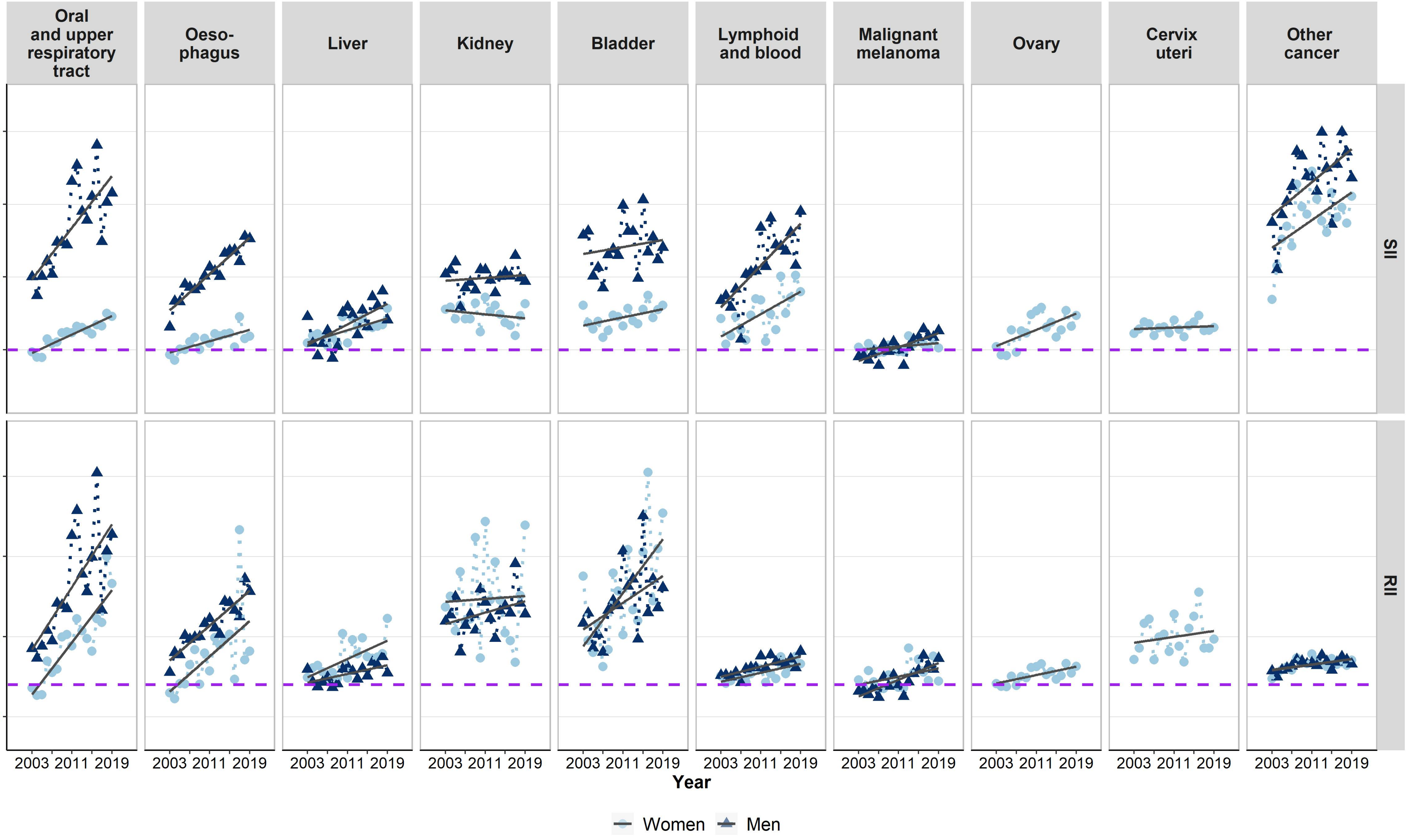


**Note: SII, Slope Index of Inequality; RII, Relative Index of Inequality**
